# Self-discontinuation of Pre-Exposure Prophylaxis Among Adolescent Girls and Young Women in Mazowe District, Zimbabwe

**DOI:** 10.1101/2025.11.06.25339707

**Authors:** Godwin Choga, Owen Mugurungi, Gerald Shambira, Addmore Chadambuka, Tsitsi Patience Juru, Notion Gombe, Mufuta Tshimanga, Richard Makurumidze

**Author notes:** Corresponding Author: Godwin Choga.

## Abstract

**Background:** Adolescent girls and young women (AGYW) in Zimbabwe are at high risk of acquiring HIV. In 2020, HIV incidence among AGYW was 9.5 times that of males. AGYW are targeted for Pre-Exposure Prophylaxis (PrEP). In Mazowe District, 28.7% of AGYW who were initiated on PrEP in 2021 self-discontinued within a month. This limits the effectiveness of PrEP and increases the risk of HIV drug resistance. We determined factors associated with PrEP self-discontinuation among AGYW in Mazowe.

**Methods:** We conducted an analytical cross-sectional study among HIV-negative AGYW initiated on PrEP. Participants were randomly selected proportionate to number initiated on PrEP in 20 health facilities in Mazowe District. Using Interviewer-administered questionnaires, data was collected on sexual behaviour, reasons for starting PrEP, and determinants of self-discontinuation. We performed logistic regression analysis at 5% significance-level to determine independent factors for PrEP self-discontinuation using Epi Info 7.2.

**Results:** Of the 384 AGYW included in the study, 57.8% were married, 24.5% were divorced, and 17.7% were never married. Commonly reported reasons for starting PrEP were multiple sexual partners (29.2%) and promiscuous sexual partners (29.2%). The proportion of PrEP self-discontinuation was 41% with a median time to self-discontinuation of 4 (IQR 2-5) months. Of 157 AGYW who self-discontinued PrEP, 59.2% had multiple sexual partners, and 57.3% had transactional sex after discontinuing PrEP. Independent factors associated with PrEP self-discontinuation among AGYW were being a sex worker [aOR 4.86; 95% CI (1.33-17.69)], non-disclosure of PrEP status to sexual partner [aOR 3.63; 95% CI (2.13-6.19)], being discouraged to take PrEP by sexual partner [aOR 3.04; 95% CI (1.31-7.04)] and experiencing PrEP side effects [aOR 2.38; 95% CI (1.49-3.81)].

**Conclusions:** Being a sex worker, lack of partner support, and PrEP side effects were significant risk factors for self-discontinuation of PrEP. Improved, differentiated PrEP services’ provision and investing in community awareness may enhance PrEP retention.

## Introduction

Significant strides have been made to end HIV and AIDS as a global pandemic. According to the 2025 Global AIDS update report, new HIV infections have been reduced by 40% from the 2010 statistics, and the reduction was largely in Eastern and Southern Africa, where a reduction of 56% was recorded [1]. Despite the progress made against HIV globally, adolescents and young women (AGYW) continue to be at greater risk of acquiring new infections. Globally, nearly 1000 AGYW are infected with HIV daily [2]. In sub-Saharan Africa, AGYW constitutes about 10% of the population, but in the year 2024, 28% of incident cases of HIV were amongst this subpopulation [1]. In Zimbabwe, AGYW are also disproportionately affected by new HIV infections. The most recent, nationally representative data for Zimbabwe showed that the incidence of HIV among AGYW was 0,76% whilst that for the adolescent boys and young men was 0.08% [3]. About 54.2% of young people in Zimbabwe who reported that they had sex within the year prior to the survey had sex with a nonmarital, non-cohabitating partner. The proportion of those who did not use a condom when they last had sex was 37.3% [3]. Social isolation, poverty, and discriminatory cultural norms contribute to vulnerability to HIV infections. The economically disadvantaged AGWY are usually unable to negotiate for safe sex [2,4,5].

To reduce new HIV infections, the World Health Organization recommends a Combination HIV Prevention approach. Combination HIV Prevention, as defined by UNAIDS, is “rights-based, evidence-informed, and community-based programs that promote a combination of biomedical, behavioural, and structural interventions designed to meet the HIV prevention needs of specific people and communities” [6]. Pre-Exposure prophylaxis (PrEP) is one of the biomedical interventions for HIV prevention. PrEP is the use of antiretroviral medicines in HIV negative people to prevent the acquisition of HIV, and when used consistently, it reduces the risk of HIV infection by over 90% [7]. However, the protection offered by PrEP depends on the proper use of the medication. A client wishing to discontinue PrEP should consult their healthcare provider so that a risk assessment can be done and a safe weaning process can be initiated [8]. PrEP should be stopped when the period of substantial HIV risk has lapsed. An HIV test should be done and documented at the time of stopping PrEP. The client should be offered counselling on other HIV prevention methods and be advised to return to the facility if they wish to restart PrEP. Failure to wean off PrEP medicines in the correct way results in an increased risk of contracting HIV [9,10].

As part of combination HIV prevention, Zimbabwe included Pre-Exposure Prophylaxis (PrEP) for high-risk groups in the national antiretroviral therapy guidelines in the year 2016 [11]. A phased approach to the provision of PrEP services was implemented, starting with Wilkins Infectious Disease Hospital in the capital city of Harare and four districts, which were Mutare, Chipinge, Gweru, and Bulawayo. The lessons learned from the initial PrEP implementation phase guided the rollout of PrEP services to other public health institutions in Zimbabwe [12]. Mazowe District, as one of the high-burden districts concerning new HIV infections among AGYW, was included in the rollout of PrEP services in 2019.

The Mazowe District HIV program data for the year 2021 revealed that 28.7% of AGYW initiated on PrEP in Mazowe District self-discontinued PrEP within a month of initiation. This reflects a high proportion of AGYW who discontinue PrEP before their periods of high-risk lapse. Self-discontinuation of PrEP results in the AGYW not being fully protected against HIV infection during their periods of high-risk and lost opportunities for getting counselling on other methods of HIV prevention, therefore increasing their risk of acquiring HIV [9,10]. To our knowledge, there are limited studies conducted on the self-discontinuation of PrEP among the AGYW, and those found in the literature are mostly qualitative studies. We therefore studied the quantitative determinants of self-discontinuation of PrEP in Mazowe District in 2021 to improve HIV prevention programming.

## Methods

### Study Setting

The study was conducted at health facilities that were offering PrEP in Mazowe District, in Mashonaland Central Province of Zimbabwe. Mazowe District had 34 health facilities which offered PrEP services, comprising three hospitals, one rural hospital, and 30 clinics. Mazowe District had a population of 293 362 people, and the HIV prevalence was 18.6% [12,13]. The economic activities in Mazowe District were mainly farming and mining.

### Study Design

An analytical cross-sectional study was conducted in Mazowe District. An analytical cross-sectional study was used as it allowed for the determination of the proportion of PrEP self-discontinuation among AGYW in Mazowe District at the time of conducting the study. There were limited resources for conducting the study, so an analytical cross-sectional study was ideal as it was relatively cheap and less time-consuming. Furthermore, the study design would provide evidence to inform PrEP public health planning by providing a snapshot of the status of PrEP programming for AGYW in Mazowe District.

### Study participants

The study participants comprised HIV negative adolescent girls and young women who were initiated on PrEP at health facilities in Mazowe District in the year 2021. Adolescent girls and young women aged between 16 and 24 years who were initiated on PrEP at health facilities in Mazowe District in the year 2021 were included in the study. Girls who were below the age of 16 years were excluded from the study as they were below the age of sexual consent in Zimbabwe.

### Sampling procedure

Using Dobson formula: n = Z_a_^2^ (p) (1-p)/delta^2^, where Z_a_=1.96, p=0.5 to optimise the sample size, and delta is 0.05, a sample size of 384 study participants was reached. A total of twenty health facilities were randomly selected from the 34 health facilities offering PrEP in Mazowe District. From the PrEP register, the AGYW initiated on PrEP in the year 2021 were identified and assigned numbers. The number of AGYWs to be recruited at each facility was calculated according to the proportion of AGYWs that were initiated on PrEP at that facility in the year 2021. Random selection of the AGYWs to be interviewed at each facility was done using an electronic random number generator.

### Study variables

The outcome variable was PrEP self-discontinuation, which was defined as any adolescent girl or young woman who was initiated on PrEP in Mazowe District in 2021 and discontinued PrEP without advice from a health worker. The other variables that were collected during interviews with the AGYW were demographic characteristics of the AGYW, reasons for starting PrEP, sexual behaviour after self-discontinuing PrEP, time to PrEP self-discontinuation, and the determinants of PrEP discontinuation

### Data Collection Tool and procedure

An interviewer-administered questionnaire was used to collect data from the AGYW. The questionnaire was validated by being reviewed by experts from the National AIDS and TB Program in the Ministry of Health and Child Care. The questionnaire was pre-tested at three health facilities in Mazowe District, which were not included in the study. Data collection for the study was conducted by the lead author from August to September 2022.

### Data Analysis

The data collected was cleaned and captured using the Epi Info 7.2 statistical package. The Epi Info 7.2 software was used to conduct univariate and bivariate analysis to generate frequencies, proportions, prevalence Odds Ratios (pORs), corresponding 95% Confidence Interval (CI), and p-values. Backward stepwise logistic regression analysis was conducted to determine the independent determinants of PrEP self-discontinuation among the AGYW. All variables associated with PrEP self-discontinuation with a p-value ≤ 0.25 were included in the logistic regression model. All variables with a p-value <0.05 were statistically significant.

### Permission and Ethical Considerations

Permission to carry out the study was obtained from the Provincial Medical Director for Mashonaland Central Province, the District Medical Officer for Mazowe District, and the Ministry of Health and Child Care, Health Studies Office. Ethical approval was sought from the University of Zimbabwe, Faculty of Medicine and Health Sciences and Parirenyatwa Hospital Joint Research Ethics Committee (JREC/305/2022), and the Health Studies Office. Written informed consent was obtained from the AGYW who were initiated on PrEP in 2021 and were recruited into this study. Confidentiality was assured and maintained throughout the study by interviewing each participant privately and ensuring that no information obtained was disclosed to any other person. Names of participants were not written on the questionnaires. All questionnaires are being kept under lock and key.

## Results

### Demographic characteristics of study participants

A total of 384 adolescent girls and young women were interviewed, with 285 (74.2%) of them being in the 20-25 years age group. The median age was 22 years [Interquartile Range (IQR):19-23]. Of the 384 AGYW, 222 (57.8%) were married, 94 (24.5%) were divorced, and 68 (17.7%) were never married. Nine (2.4%) of the AGYW had never been enrolled in school, whilst 124 (32.3%) reached primary school level, 245 (63.8%) reached secondary school level, and six (1.5%) reached the tertiary education level. The major sources of income for the AGYW were informal employment (32.3%), sex work (20.1%), self-employment (11.9%), and formal employment (3.7%), whilst 32% of the AGYW had no source of income of their own. The majority (82%) of the AGYW were Christians (Table 1).

**Table 1:** Demographic characteristics of study participants in Mazowe District. 2021.

| Variable | PrEP self-discontinuation |  |
| --- | --- | --- |
|  | Yes n = 157 (%) | No n = 227 (%) |
| Age |  |  |
| 16-19 years | 42 (26.7) | 57 (25.1) |
| 20-24 years | 115 (73.3) | 170 (74.9) |
| Median age (Q1; Q3) | 22 (19; 23) | 22 (19; 23) |
| Marital status |  |  |
| Married | 69 (44.0) | 153 (67.4) |
| Never married | 36 (22.9) | 32 (14.1) |
| Divorced | 52 (33.1) | 42 (18.5) |
| Level of education |  |  |
| Primary | 61 (38.9) | 63 (27.8) |
| Secondary | 89 (56.7) | 156 (68.7) |
| Tertiary | 4 (2.5) | 2 (0.9) |
| None | 3 (1.9) | 6 (2.6) |
| Occupation |  |  |
| Formally employed | 5 (3.2) | 9 (4.0) |
| Informally employed | 53 (33.7) | 117 (51.5) |
| Sex worker | 56 (35.7) | 21 (9.3) |
| None | 43 (27.4) | 80 (35.2) |
| Religion |  |  |
| Christianity | 130 (82.8) | 185 (81.5) |
| African Traditional Religion | 2 (1.3) | 3 (1.3) |
| Islam | 2 (1.3) | 6 (2.7) |
| None | 23 (14.6) | 33 (14.5) |

### Reported reasons for starting PrEP by AGYW in Mazowe District

Having multiple partners, 112/384 (29.2%) and knowing that a sexual partner was promiscuous, 112/384 (29.2%), were the major reasons for AGYW to start PrEP. The other reasons for starting PrEP were being in a long-distance sexual relationship 40/384 (10.4%), having a sexual partner whose HIV status was unknown 39/384 (10.2%), a history of developing a sexually transmitted illness 34/384 (8.9%), and being in an HIV sero-discordant relationship 29/384 (7.6%).

### PrEP medicine side effects

Of the 384 respondents, 176 (45.8%) reported they had experienced side effects whilst taking PrEP medications. The reported side effects by the AGYW were nausea and vomiting 39/176 (22.2%), dizziness 35/176 (19.9%), headache 30/176 (17.1%), diarrhoea 25/176 (14.2%), generalised body weakness 25/176 (14.2%), having an excessive appetite 13/176 (7.4%) and skin rash 9/176 (5.0%).

### Proportion of PrEP self-discontinuation among AGYW in Mazowe District

Of the 384 AGYW enrolled in the study, 157(40.9%) reported that they self-discontinued PrEP. The median time to PrEP self-discontinuation by the AGYW was four months, with a 25^th^ percentile of two months and a 75^th^ percentile of five months.

### Sexual behaviour of the AGYW who self-discontinued PrEP

Ninety-five (60.5%) of the 157 AGYW who self-discontinued PrEP reported that they had unprotected sexual intercourse, whilst 93 (59.2%) had multiple sexual partners and 90 (57.3%) had been involved in transactional sex in the six months preceding the interviews. There were 82 AGYW (52.2%) who reported that they had a partner with an unknown HIV status, and six (3.8%) had an HIV sero-discordant sexual partner. (Table 2).

**Table 2:**
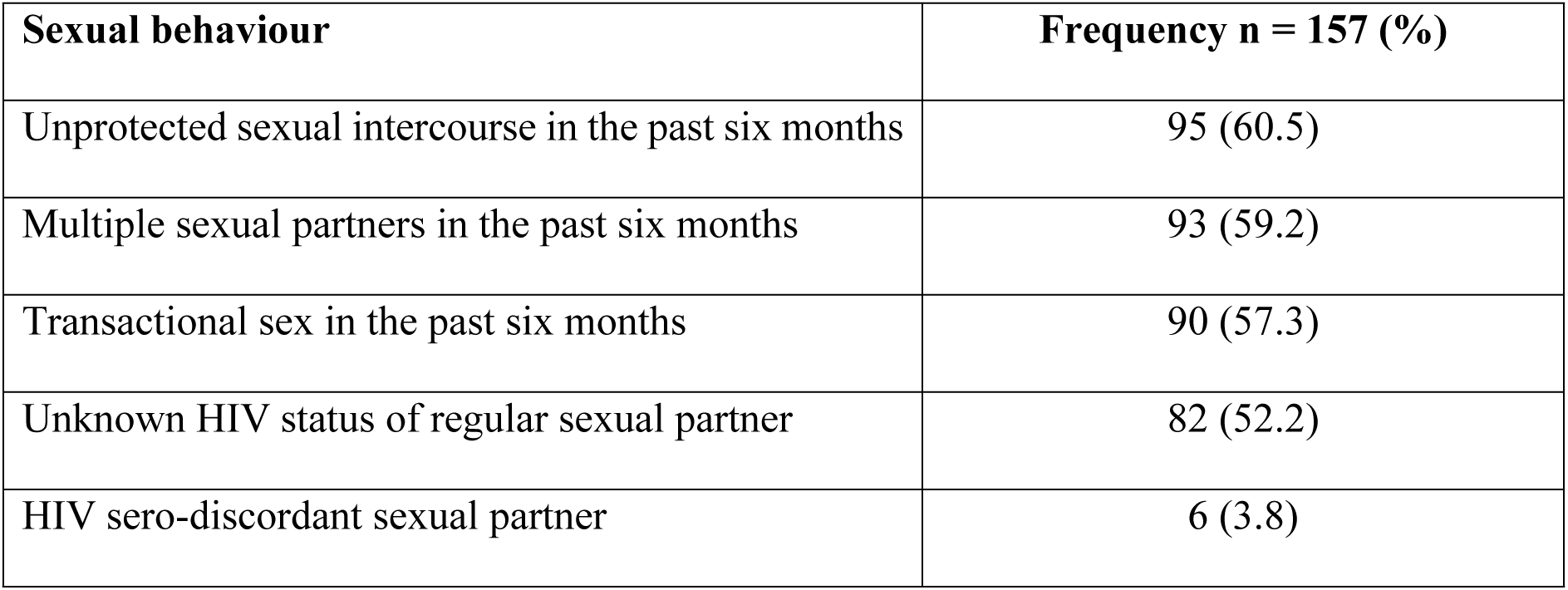
Sexual behaviour of the AGYW who self-discontinued PrEP, Mazowe District, 2021.

| Sexual behaviour | Frequency n = 157 (%) |
| --- | --- |
| Unprotected sexual intercourse in the past six months | 95 (60.5) |
| Multiple sexual partners in the past six months | 93 (59.2) |
| Transactional sex in the past six months | 90 (57.3) |
| Unknown HIV status of regular sexual partner | 82 (52.2) |
| HIV sero-discordant sexual partner | 6 (3.8) |

### Factors Associated with PrEP Self-discontinuation among AGYW

On bivariate analysis the variables which had a p-value less than 0.25 which were included in the multivariate analysis were marital status, main source of income, experiencing PrEP medicine side effects, partner discouragement, alcohol use, transport cost, average waiting time at health facility, unsatisfactory PrEP clinic waiting area and health worker attitude. After controlling for confounding, the independent factors associated with PrEP self-discontinuation among the AGYW were being a sex worker [adjusted pOR 4.86. 95% CI (1.33-17.69)], experiencing PrEP medicine side effects [adjusted pOR 2.38, 95% CI (1.49-3.81)], experiencing partner discouragement [adjusted pOR 3.04, 95% CI (1.31-7.04)], and lack of disclosure to partner [adjusted pOR 3.63, 95% CI (2.13-6.19)]. (Table 3).

**Table 3:** Factors associated with PrEP self-discontinuation among AGYW, Mazowe District, 2021.

| Variable | PrEP self-discontinuation |  | pOR (95% CI) | P value | Adjusted pOR (95% CI) | P value |
| --- | --- | --- | --- | --- | --- | --- |
|  | Yes n (%) | No n (%) |  |  |  |  |
| Age |  |  |  |  |  |  |
| 16-19 years | 42 (26.7) | 57 (25.1) | 1.08 (0.68-1.72) | 0.737 |  |  |
| 20-24 years | 115 (73.3) | 170 (74.9) |  |  |  |  |
| Married |  |  |  |  |  |  |
| Yes | 69 (44.0) | 153 (67.4) | 0.38 (0.25-0.58) | **<0.001** |  |  |
| No | 88 (56.0) | 74 (32.6) |  |  |  |  |
| Main source of income |  |  |  |  |  |  |
| Formal employment | 5(3.2) | 9(4.0) | 1 |  | 1 |  |
| Informal employment | 46(29.3) | 78(34.4) | 1.06(0.34-3.36) | 0.919 | 1.73(0.50-5.98) | 0.386 |
| None | 43(27.4) | 80(35.2) | 0.98(0.31-3.07) | 0.955 | 1.52(0.44-5.29) | 0.955 |
| Self employed | 7(4.5) | 39(17.2) | 0.32(0.08-1.26) | 0.103 | 0.62(0.15-2.61) | 0.103 |
| Sex work | 56(35.6) | 21(9.2) | 4.80(1.44-15.98) | **0.011** | 4.86(1.33-17.69) | **0.017** |
| Experienced PrEP medicine side effects |  |  |  |  |  |  |
| Yes | 97 (61.8) | 85 (37.4) | 2.70 (1.78-4.11) | **<0.001** | 2.38(1.49-3.81) | **<0.001** |
| No | 60 (38.2) | 142 (62.6) |  |  |  |  |
| Experienced stigma associated PrEP |  |  |  |  |  |  |
| Yes | 47 (29.9) | 62 (27.3) | 1.14 (0.73-1.78) | 0.575 |  |  |
| No | 110 (70.1) | 165 (72.7) |  |  |  |  |
| Partner discouragement |  |  |  |  |  |  |
| No | 71(45.2) | 180(79.3) | 1 |  | 1 |  |
| Yes | 17(10.8) | 12(5.3) | 3.59(1.63-7.90) | **0.001** | 3.04(1.31-7.04) | **0.009** |
| Not disclosed | 69(44.0) | 35(15.4) | 5.00(3.06-8.16) | **<0.001** | 3.63(2.13-6.19) | **<0.001** |
| Alcohol use |  |  |  |  |  |  |
| Yes | 34 (21.7) | 36 (15.9) | 1.47 (0.87-2.47) | 0.148 |  |  |
| No | 123 (78.3) | 191 (84.1) |  |  |  |  |
| Negative health worker attitude |  |  |  |  |  |  |
| Yes | 15 (9.6) | 10 (4.4) | 2.29 (1.00-5.24) | **0.044** |  |  |
| No | 142 (90.4) | 217 (95.6) |  |  |  |  |
| Unsatisfactory PrEP Clinic Waiting Area |  |  |  |  |  |  |
| Yes | 28(17.8) | 30 (13.2) | 1.43 (0.81-2.50) | 0.216 |  |  |
| No | 129 (82.2) | 197 (86.8) |  |  |  |  |
| Average waiting time at health facility |  |  |  |  |  |  |
| ≤60 mins | 102(65.0) | 186(81.9) | 1 |  |  |  |
| 61-120 mins | 29(18.5) | 18(7.9) | 2.94(1.56-5.55) | **0.001** |  |  |
| >120 mins | 26(16.5) | 23(10.2) | 2.06(1.12-3.80) | **0.020** |  |  |
| Transport cost to health facility |  |  |  |  |  |  |
| Yes | 23(14.7) | 53(23.4) | 0.56(0.33-0.97) | **0.035** |  |  |
| No | 134(85.3) | 174(76.6) |  |  |  |  |
| Distance from health facility |  |  |  |  |  |  |
| ≤5 km | 114 (72.6) | 167 (73.6) | 0.95 (0.60-1.51) | 0.835 |  |  |
| >5km | 43 (27.4) | 60 (26.4) |  |  |  |  |

## Discussion

The study revealed a high prevalence of PrEP self-discontinuation among adolescent girls and young women in Mazowe District. Being a sex worker, experiencing PrEP medicine side effects, lack of disclosure of PrEP status, and facing partner discouragement were associated with PrEP self-discontinuation among AGYW. The majority of the AGYW who self-discontinued PrEP reported having engaged in unprotected sex, transactional sex, and having multiple sexual partners within the six months preceding the interviews.

Sex workers are one of the marginalised groups in terms of access to health care services. The previous laws, which criminalise sex work and the fear of being judged by health care workers, serve as barriers to accessing health services by sex workers [14,15]. Sex workers are also very migratory, moving to areas where they are likely to meet clients who can pay for their services [16]. There are gold panning activities in Mazowe District, and the sex workers usually migrate from their usual residential areas to these gold panning areas. There is a possibility that the refill times for PrEP may lapse before they return to their usual places of residence. Their engagement in sexual intercourse with multiple partners, transactional sex, having sexual encounters with people of unknown HIV status, and increased chances of sexual violence place these sex workers at high risk of acquiring HIV [17,18]. PrEP self-discontinuation among the sex workers is therefore a cause for concern, and strategies should be put in place to retain them on PrEP. Innovative and differentiated models of PrEP service delivery could be useful approaches to this key population.

Experiencing medicine side effects negatively affected the continuation of AGYW on PrEP. The commonest side effects reported by the AGYW were nausea and vomiting, dizziness, and headache. These were consistent with the documented side effects of Tenofovir and Emtricitabine, which constitute the combination of drugs used for PrEP [19]. AGYW who take PrEP medicines are usually not ill and will be taking the medicines for prevention rather than for treatment. It could be difficult for the AGYW to take medicines that cause them discomfort when they are not suffering from any illness. Similar findings were revealed by studies by I Beesham et al in South Africa, and U Koppe et al in Germany [20,21]. Most of the PrEP side effects occur during, but not limited to, the early days of initiation and usually subside without the client stopping medications within a timeframe of a week to a month [22]. Enhanced counselling on management of PrEP side effects could reduce the self-discontinuation rates among the AGYW. The implication of AGYW self-discontinuing PrEP due to transient and self-limiting medicine side effects is severe, as this increases their risk of HIV transmission.

Adolescent girls and young women who were discouraged from taking PrEP by their partners and those who did not disclose their PrEP status to their partners were more likely to self-discontinue PrEP compared to those who had encouraging partners. This could be due to a lack of support in continuing and adhering to PrEP medicines. Fear of verbal or physical abuse and possibly fear of rejection from their partners could be leading AGYW not to disclose that they are taking PrEP. There are usually misconceptions that PrEP medicines increase a person’s chances of being promiscuous. Some perceive these medicines as treatment for HIV, and they label PrEP users as people living with HIV. This increases the chances of self-discontinuation of PrEP medicines by the AGYW. However, studies have shown that disclosing the PrEP status to partners, family, or friends results in improved support in the form of reminders to take medicines and providing morale support during periods of demotivation [21,23].

No association was found between health system-related determinants and self-discontinuation of PrEP among the AGYW in Mazowe District. This could be due to task sharing on providing counseling sessions at the health facility between the nurses, Primary Counsellors, and a trained peer leader known as PrEP champion, who is one of the AGYW on PrEP. This reduces the workload on the nurses and improves the waiting time at the health facility. This was consistent with findings by Rutstein et al, who found that the majority of the PrEP discontinuations were likely due to client factors [24]. This implies that, to address new HIV infections among the AGYW, there should be focused discussions on addressing client-related factors associated with PrEP self-discontinuation.

The majority of AGYW who self-discontinued PrEP were involved in sexual behaviours that predisposed them to acquiring HIV in the six months preceding the interviews. The activities were unprotected sexual intercourse, having multiple sexual partners, engaging in transactional sex, and having sexual relations with people with unknown HIV status. The high proportion of AGYW who never reached the secondary school level could be a proxy indicator for poverty in Mazowe District. This poverty could also be driving the risk behaviours by the AGYW, such as engaging in transactional sex. The findings are consistent with findings by U Koppe et al, who showed inconsistent condom use and having multiple sexual partners among former PrEP users [20]. Engaging in activities such as having transactional sex, having unprotected sexual intercourse, and multiple sexual partners after discontinuing the highly efficacious PrEP predisposes AGYW to new HIV infections.

## Limitations

There is a possibility of a social desirability bias in the study. Sexuality issues are sensitive, especially among the AGYW population. Furthermore, there were married women recruited in the study, and they might shy away from telling the truth about their actual sexual behaviours. We tried to mitigate for that by conducting the interviews in privacy and reassuring the participants that no identifying information would be collected and that their information would be kept confidential.

The interviewer was a healthcare worker, and the participants could possibly think that there was a possibility of the interviewer divulging their responses to the health facility staff. This could potentially result in an underestimation of the association between health system-related factors and PrEP self-discontinuation. We tried to account for this by conducting the interviews outside the health facility premises and reassuring the respondents that the interviewer did not work in Mazowe District and that their information would be kept confidential.

## Conclusion

The independent factors associated with PrEP self-discontinuation among the AGYW were being a sex worker, experiencing PrEP medicine side effects, partner discouragement, and lack of disclosure to the partner. There was no association between health system-related determinants, such as health worker attitude and waiting time at the health facility, and self-discontinuation of PrEP. The AGYW who self-discontinued PrEP engaged in sexual behaviours that predispose them to acquiring HIV within six months prior to the interviews.

We recommend the introduction of PrEP service delivery models that are tailor-made to the needs of individual PrEP users, such as community PrEP refill groups and fast-track refill at health facilities. There should be enhanced counselling on the management of PrEP medicine side effects and the importance of disclosure of PrEP status to sexual partners. Community awareness campaigns to demystify PrEP might go a long way in making people understand the PrEP program and for them to accept when their partners decide to take PrEP. The government should embark on economic empowerment projects for AGYW to reduce sex work among this population.

## Data Availability

The file is publicly available on Figshare. The link to the file is https://figshare.com/articles/dataset/Self-Discontinuation_of_Pre-Exposure_Prophylaxis_among_Adolescent_Girls_and_Young_Women_Mazowe_Zimbabwe_2021_/30418666?file=58961998

## Acknowledgements

We would like to express our sincere gratitude to the staff in the Faculty of Medicine and Health Sciences, Department of Global, Public Health and Family Medicine, and the Health Studies Office for the guidance and support that they rendered. Many thanks go to the Mazowe District Health Executive, the Mashonaland Central Provincial Health Executive, Bridget Chiururwi, the Program Officer for Pangea AIDS Trust (PZAT), and the PrEP champions supported by PZAT for their assistance in the identification and mobilisation of the sampled AGYW for the interviews, and all the participants for their consent to be part of the study.

## Declarations

### Authors’ contributions

**Conceptualization:** Godwin Choga, Mufuta Tshimanga, Owen Mugurungi, Tsitsi Patience Juru, Notion Gombe, Gerald Shambira, Addmore Chadambuka, Richard Makurumidze.

**Data curation:** Godwin Choga, Mufuta Tshimanga, Gerald Shambira, Owen Mugurungi, Tsitsi Patience Juru.

**Formal analysis:** Godwin Choga, Mufuta Tshimanga, Tsitsi Patience Juru, Gerald Shambira, Addmore Chadambuka, Notion Gombe, Richard Makurumidze.

**Investigation:** Godwin Choga, Owen Mugurungi.

**Methodology:** Godwin Choga, Mufuta Tshimanga, Owen Mugurungi, Tsitsi Patience Juru, Notion Gombe, Gerald Shambira, Addmore Chadambuka, Richard Makurumidze.

**Project administration:** Godwin Choga, Mufuta Tshimanga, Owen Mugurungi, Tsitsi Patience Juru, Notion Gombe, Gerald Shambira, Richard Makurumidze.

**Resources:** Godwin Choga, Mufuta Tshimanga, Owen Mugurungi, Gerald Shambira, Tsitsi Patience Juru.

**Supervision:** Mufuta Tshimanga, Owen Mugurungi, Tsitsi Patience Juru, Notion Gombe, Gerald Shambira, Addmore Chadambuka, Richard Makurumidze.

**Validation:** Mufuta Tshimanga, Owen Mugurungi, Gerald Shambira, Richard Makurumidze.

**Writing – original draft:** Godwin Choga, Mufuta Tshimanga, Owen Mugurungi, Tsitsi Patience Juru, Notion Gombe, Gerald Shambira, Addmore Chadambuka.

**Writing – review & editing:** Godwin Choga, Mufuta Tshimanga, Owen Mugurungi, Tsitsi Patience Juru, Notion Gombe, Gerald Shambira, Addmore Chadambuka, Richard Makurumidze.

### Permission to publish

Permission to publish the article was obtained.

### Ethical consideration

All the study participants signed informed consent forms prior to participation in the study. The names of the study participants were not written on the data collection tools.

